# Artificial Intelligence in Healthcare: 2023 Year in Review

**DOI:** 10.1101/2024.02.28.24303482

**Authors:** Raghav Awasthi, Shreya Mishra, Rachel Grasfield, Julia Maslinski, Dwarikanath Mahapatra, Jacek B. Cywinski, Ashish K. Khanna, Kamal Maheshwari, Chintan Dave, Avneesh Khare, Francis A. Papay, Piyush Mathur

## Abstract

**Background:** The infodemic we are experiencing with AI related publications in healthcare is unparalleled. The excitement and fear surrounding the adoption of rapidly evolving AI in healthcare applications pose a real challenge. Collaborative learning from published research is one of the best ways to understand the associated opportunities and challenges in the field. To gain a deep understanding of recent developments in this field, we have conducted a quantitative and qualitative review of AI in healthcare research articles published in 2023.

**Methods:** We performed a PubMed search using the terms, “machine learning” or “artificial intelligence” and “2023”, restricted to English language and human subject research as of December 31, 2023 on January 1, 2024. Utilizing a Deep Learning-based approach, we assessed the maturity of publications. Following this, we manually annotated the healthcare specialty, data utilized, and models employed for the identified mature articles. Subsequently, empirical data analysis was performed to elucidate trends and statistics.Similarly, we performed a search for Large Language Model(LLM) based publications for the year 2023.

**Results:** Our PubMed search yielded 23,306 articles, of which 1,612 were classified as mature. Following exclusions, 1,226 articles were selected for final analysis. Among these, the highest number of articles originated from the Imaging specialty (483), followed by Gastroenterology (86), and Ophthalmology (78). Analysis of data types revealed that image data was predominant, utilized in 75.2% of publications, followed by tabular data (12.9%) and text data (11.6%). Deep Learning models were extensively employed, constituting 59.8% of the models used. For the LLM related publications,after exclusions, 584 publications were finally classified into the 26 different healthcare specialties and used for further analysis. The utilization of Large Language Models (LLMs), is highest in general healthcare specialties, at 20.1%, followed by surgery at 8.5%.

**Conclusion:** Image based healthcare specialities such as Radiology, Gastroenterology and Cardiology have dominated the landscape of AI in healthcare research for years. In the future, we are likely to see other healthcare specialties including the education and administrative areas of healthcare be driven by the LLMs and possibly multimodal models in the next era of AI in healthcare research and publications.

## INTRODUCTION

Research and publications related to artificial intelligence (AI) in healthcare continue to grow exponentially. This exponential growth can also be seen with continued introduction of new journals focused just on AI in healthcare [1]. The research is not only related to trials exploring opportunities to apply AI in healthcare, but also related to “mature” real world testing and deployment of FDA approved AI solutions in healthcare[2,3]. The infodemic similar to what we recently saw with COVID-19 publications continues to challenge us with keeping pace with growing knowledge in this area and tease out the mature AI solutions[4,5].

The purpose of this review is to provide a comprehensive review of publications related to AI applications in healthcare for the year 2023. Both the quantity and the quality of publications continue to increase amongst all specialties. We used maturity based assessment of the publications, using a deep learning model, which performs with a high degree of accuracy[4,6,7]. Further manual analysis of data and model type to provide a detailed overview of the publications classified as mature is also provided. Unique to this year is also the significant increase in publications related to generative artificial intelligence, especially large language models (LLM), and specifically related to GPT[8]. We also provide a quantitative analysis and review of publications related to the newly evolving field of generative AI in healthcare.

## METHODOLOGY

We performed a PubMed search (Figure 1) using the terms, “machine learning” or “artificial intelligence” and “2023”, restricted to English language and human subject research as of December 31, 2023 on January 1st 2024. This search resulted in an initial pool of 23,306 publications. Our methodology has remained consistent over the past four years, which allows for comparative analysis of publications for each medical speciality, year over year[7,9].

**Figure 1:**
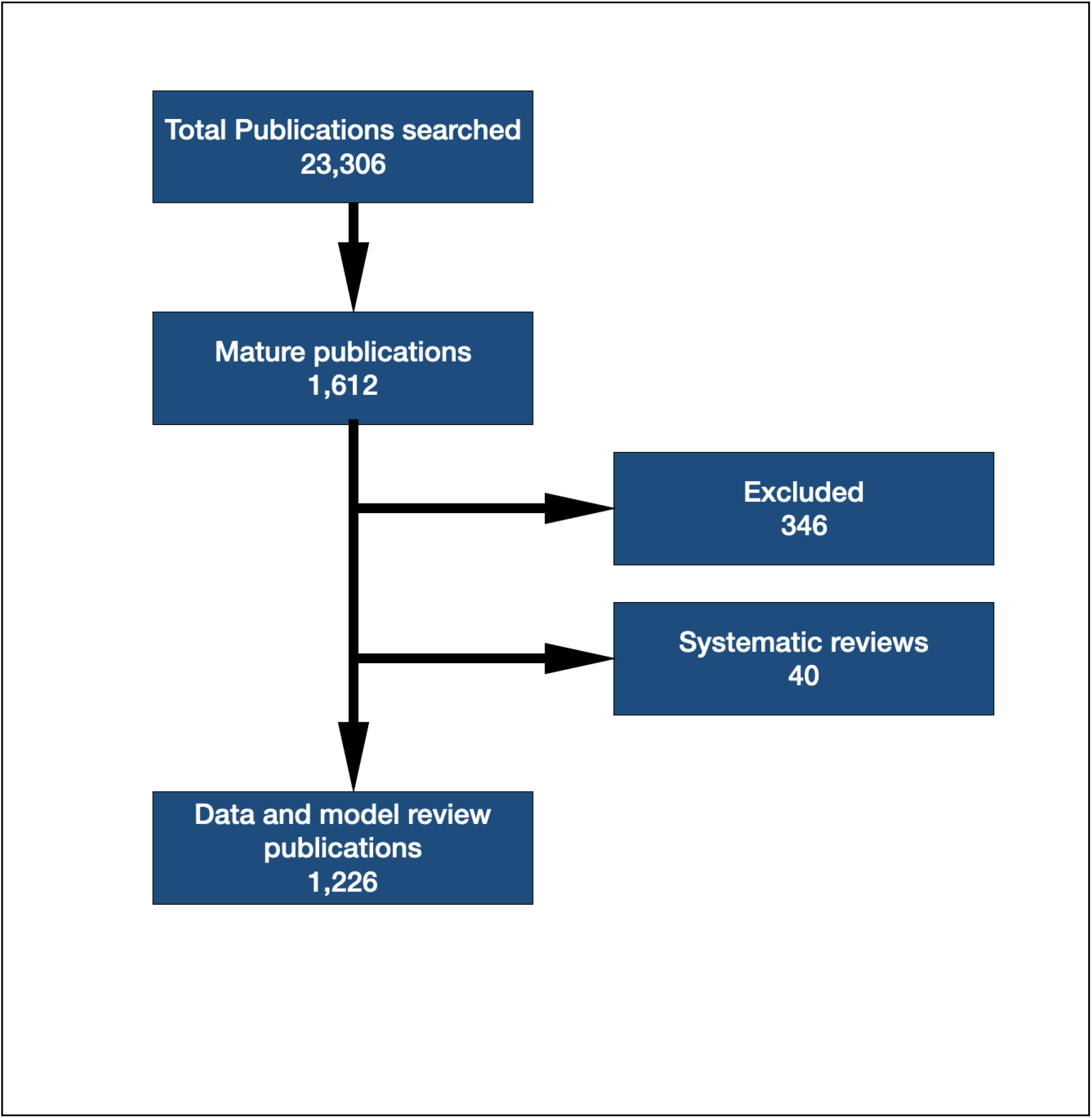
Flowchart depicting the steps involved in the inclusion and exclusion of articles. of articles

We performed qualitative evaluation of the publications’ maturity with additional details related to the type of data used and type of models developed across the healthcare spectrum. We used a Bidirectional Encoder Representation from Transformer (BERT)-based maturity classification model, that was pre-trained and validated on manually labeled data for ‘Mature’ and ‘Not Mature’ publications[6], to assess the level of maturity of each publication. The level of maturity was determined by the ability of the publication to answer the question: “Does the output of the proposed model have a direct, actionable impact on patient care by providing information to healthcare providers or automated systems?” Systematic reviews were excluded from the count of “mature” publications as they do not independently address the above question, as the models are variable in the systematic reviews[6].

Identified publications were manually reviewed and 346 publications were excluded out of the 1612 mature ones as they were not related to AI. Most of these were related to robotic surgeries or non-human studies. Further mature publications were classified based on the healthcare specialty. General category contains many of the publications related to general AI topics which were not specialty specific for instance including drug development related publications. Systematic reviews or scoping reviews were separately classified and removed prior to performing further data and model type analysis.

Finally, we manually annotated specific details from the remaining mature publications, such as data type & model type. Data type was classified manually into the four categories of data: image, text, tabular and voice. Few publications used more than one type of data for which credit was given to each of the data categories. Model type was also manually curated from the abstracts into seven different categories manually: Deep Learning (DL), Machine Learning (ML), AI General, Large Language Model (LLM), Large Vision Model (LVM), Statistical and Natural Language Processing (NLP). Those publications where the model type was not described in the abstract were placed in the AI General class. Many of these publications were related to validation studies related to proprietary AI models. Similarly, studies using statistical analysis to analyze performance of AI models were included in the statistical class.

Due to the exponential rise in LLM publications, we also performed a separate search in PubMed for LLM publications. We used “LLM” and “GPT” as keywords for the search filtering for dates from 01-01-2023 till 12-31-2023 on January 1, 2024. We also filtered our search for human and english language publications only. Our initial search yielded 808 publications, out of which 224 publications were excluded. Most of the excluded search publications were unrelated to LLMs. 584 publications were finally analyzed and classified for the 26 different specialities and used for our review.

## RESULTS

In the previous years, the total number of articles progressively increased from 3351 in 2019 to 5885 in 2020, marking a 75.59% jump, then slightly decreased to 4164 in 2021, resulting in a 29.24% decrease, and further rose to 9974 in 2022, demonstrating a significant 139.55% increase[7,9]. Also, in 2023, again a significant increase in the number of articles identified, reaching 23306. This represents a remarkable 133.7% jump **(Figure 2A)**. Also, in previous years there were 99, 250, 36, 600 mature articles in the year 2019, 2020, 2021, 2022 respectively and 2023 there were 1612 mature articles[7,9] **(Figure 2B)**.

**Figure 2:**
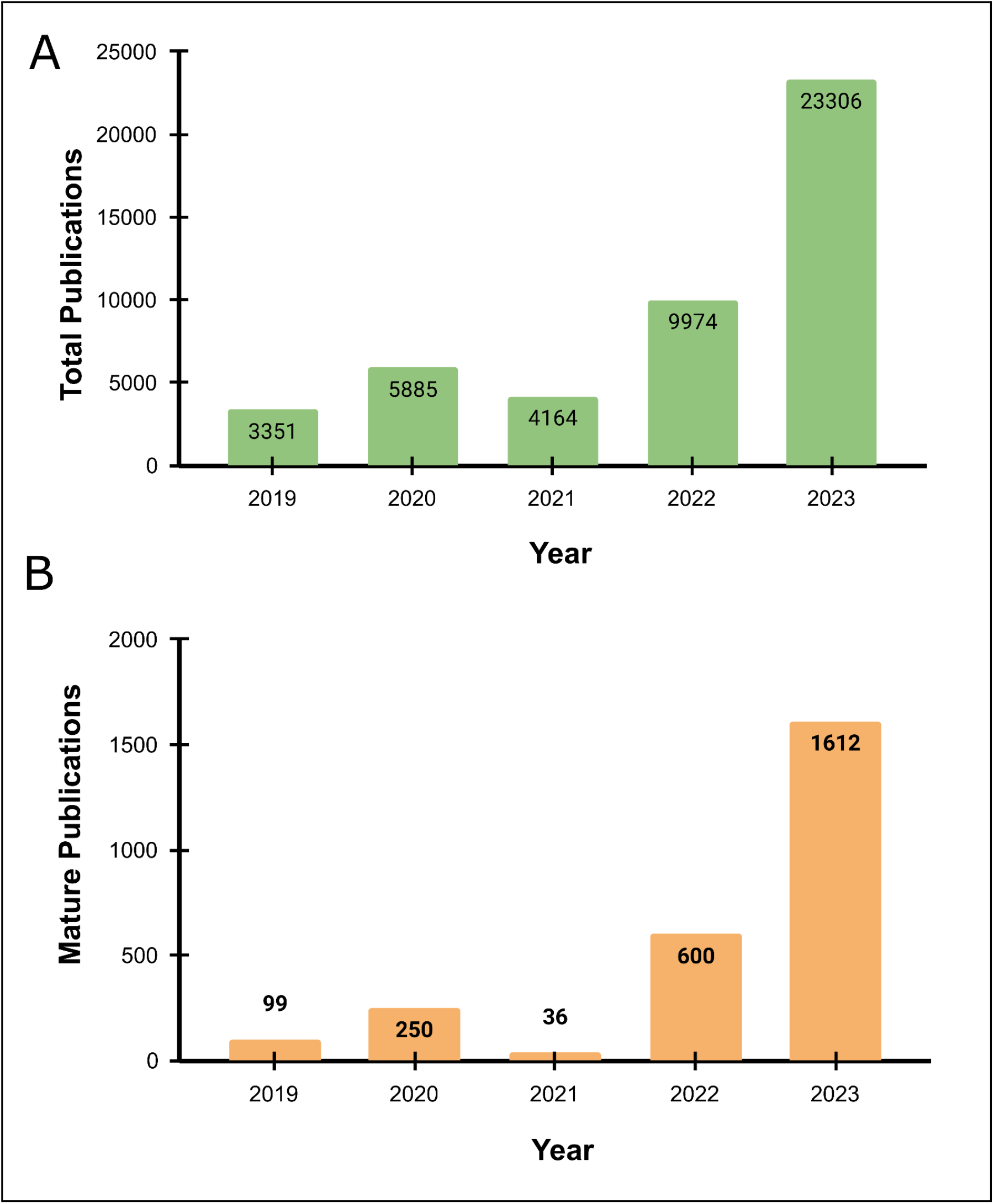
A) Bar plot showing the total number of publications from 2019-2023. B) Bar plot showing the number of mature publications from 2019-2023.

The distribution of mature articles across specialties was not evenly distributed, led by Imaging with 453 publications, followed by Gastroenterology with 86, and Ophthalmology with 78. General class of publications has 73 publications, while Pathology and Oncology both have 62 and 60 publications, respectively. Surgery and Head and Neck specialties each have 57 publications, with Cardiology following closely behind with 52 **(Table 1)**.

**Table 1:** Number of mature publications by healthcare specialty for the year 2023.

| Speciality | Number of Mature Publications |
| --- | --- |
| Imaging | 453 |
| Gastroenterology | 86 |
| Ophthalmology | 78 |
| General | 73 |
| Pathology | 62 |
| Oncology | 60 |
| Surgery | 57 |
| Head and Neck | 57 |
| Cardiology | 52 |
| Education | 48 |
| Orthopedics | 33 |
| Ob/Gyn | 21 |
| COVID-19 | 20 |
| Neurology | 19 |
| Dermatology | 18 |
| Administrative | 17 |
| Pediatrics | 16 |
| Pulmonary | 10 |
| Emergency Medicine | 8 |
| Endocrinology | 7 |
| Anesthesiology | 7 |
| Critical Care | 7 |
| Psychiatry | 5 |
| Nephrology | 4 |
| Rehabilitation Medicine | 4 |
| Genetics | 4 |

Next, we analyzed the five-year distribution of mature publications across different healthcare specialties (**Figure 3**). Imaging had the highest number of mature articles (except in 2021), with 37 in 2019, 75 in 2020, 251 in 2022, and 453 in 2023. In 2021, Gastroenterology had the highest number of mature publications, with 2, 9, 11, 24, and 86 mature articles from 2019 to 2023. Similarly, we observed a consistent increasing trend in other specialties: General had 2, 2, 0, 22, and 73 mature articles from 2019 to 2023; Surgery had 1, 3, 14, 0, and 57 from 2019 to 2023; Cardiovascular had 4, 8, 1, 33, and 52 from 2019 to 2023; Pathology had 6, 12, 0, 50, and 62 from 2019 to 2023; Head & Neck had 1, 6, 1, 24, and 57 from 2019 to 2023. Interestingly, Education had no mature articles until 2021 but saw an increase to 5 in 2022 and a substantial jump to 64 in 2023.

**Figure 3:**
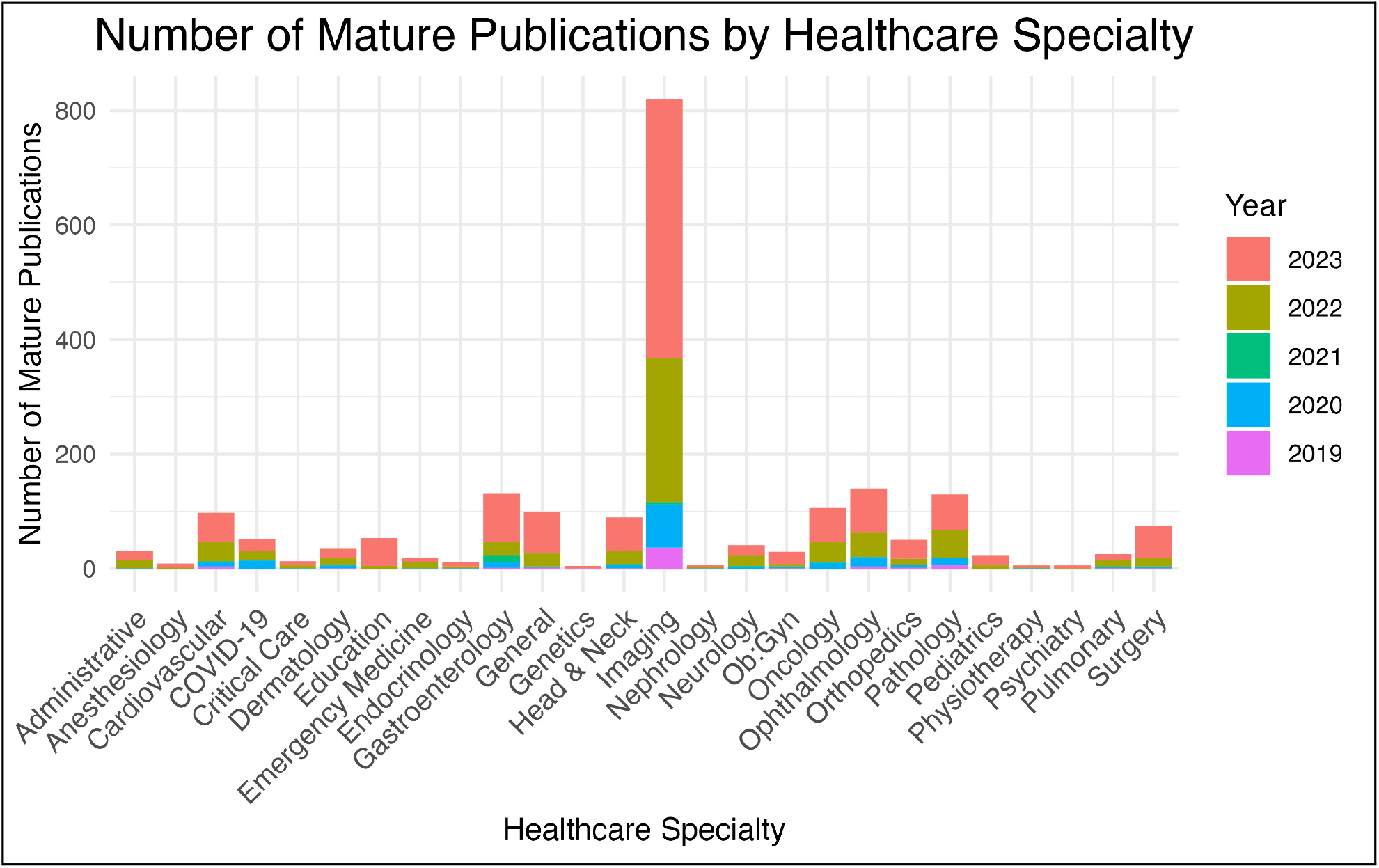
Five-Year distribution in number of mature publications in different healthcare specialties

Next, we analyzed the distribution of data types across all mature publications totaling 1612, it was evident that image data prevailed, featuring in 934 (75.2%) publications. Tabular data was utilized in 160 (12.9%) publications, followed by text data in 144 (11.6%), publications and audio data in only 4 (0.3%) publications **(Figure 4A)**. Important to note 1.4% publications used more than one type of data for which credit was given to each of the data categories. Similarly, in examining the distribution of AI/ML models employed across all publications, DL emerged as the most prominent with 730 (59.8%) publications, followed by AI General with 205 (16.8%) publications, ML (Machine Learning) with 122 (10%), LLM with 100 (8.2%), NLP with 45 (3.7%), LVM with 2 (0.4%), and Statistical approaches with 17 (1.4%) publications **(Figure 4B)**.

**Figure 4:**
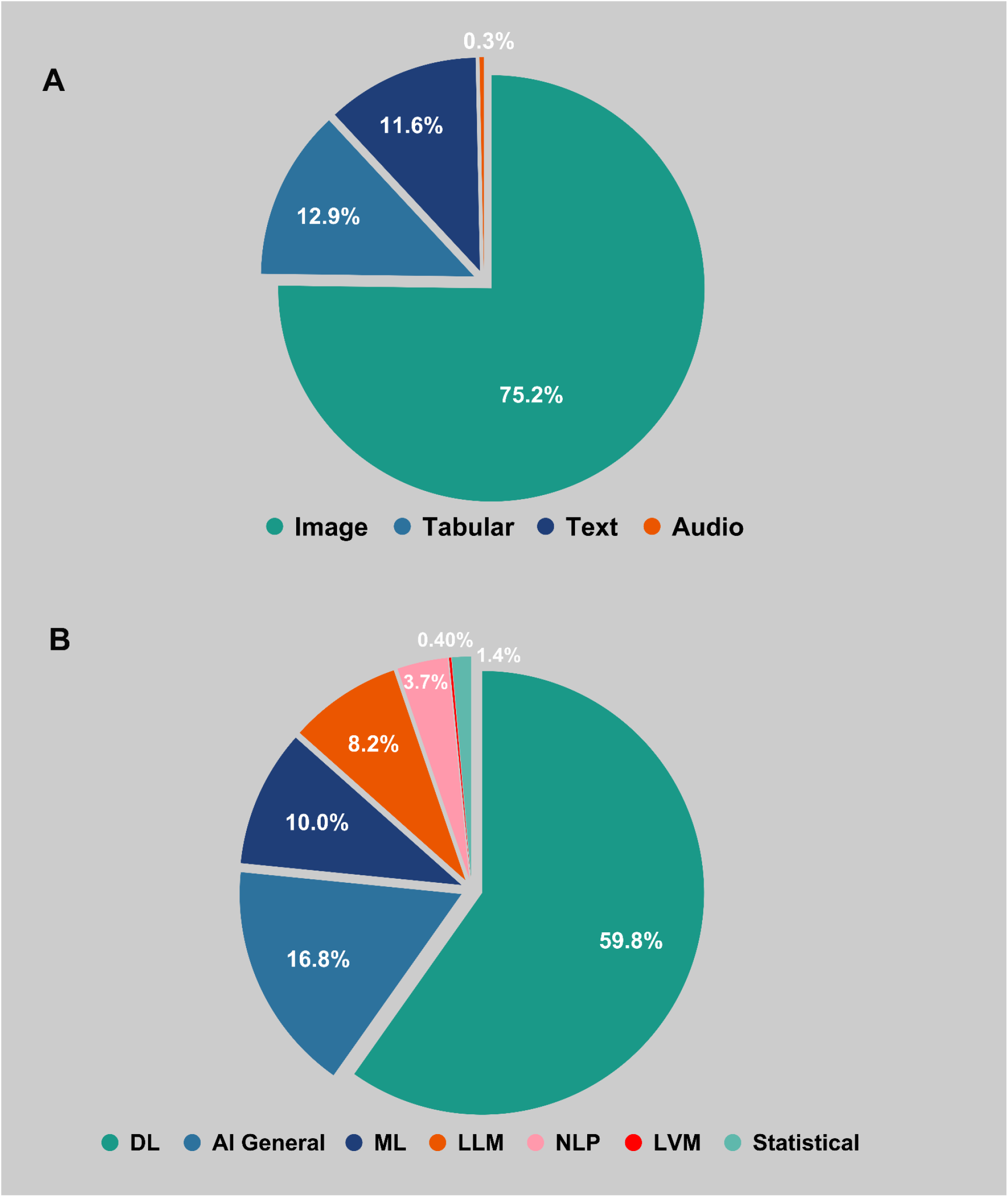
A) Distribution of data types utilized in the mature articles in the year 2023. B) Distribution of model types utilized in the mature articles in the year 2023, where DL = Deep Learning, ML = Machine Learning, LLM = Large Language Models, NLP = Natural Language Processing, LVM = Large Vision Models.

The distribution of 584 selected publications utilizing Large Language Models (LLM) across 26 various medical specialties revealed interesting findings. Overall, Imaging emerged as the specialty with the highest number of mature articles, followed by Gastroenterology and Ophthalmology. However, when considering articles incorporating LLMs, General medical topics exhibited the highest utilization, with 163 publications, followed by Surgery with 69, Education with 64, and Imaging with 55. Oncology also featured a substantial number of LLM-based publications with 26, while Ophthalmology and Psychiatry had 19 and 22, respectively. In contrast, specialties such as Gastroenterology, Pathology, and Cardiology had fewer LLM-based publications, suggesting varying levels of adoption and application of large language models across different medical research domains **(Figure 5)**.

**Figure 5:**
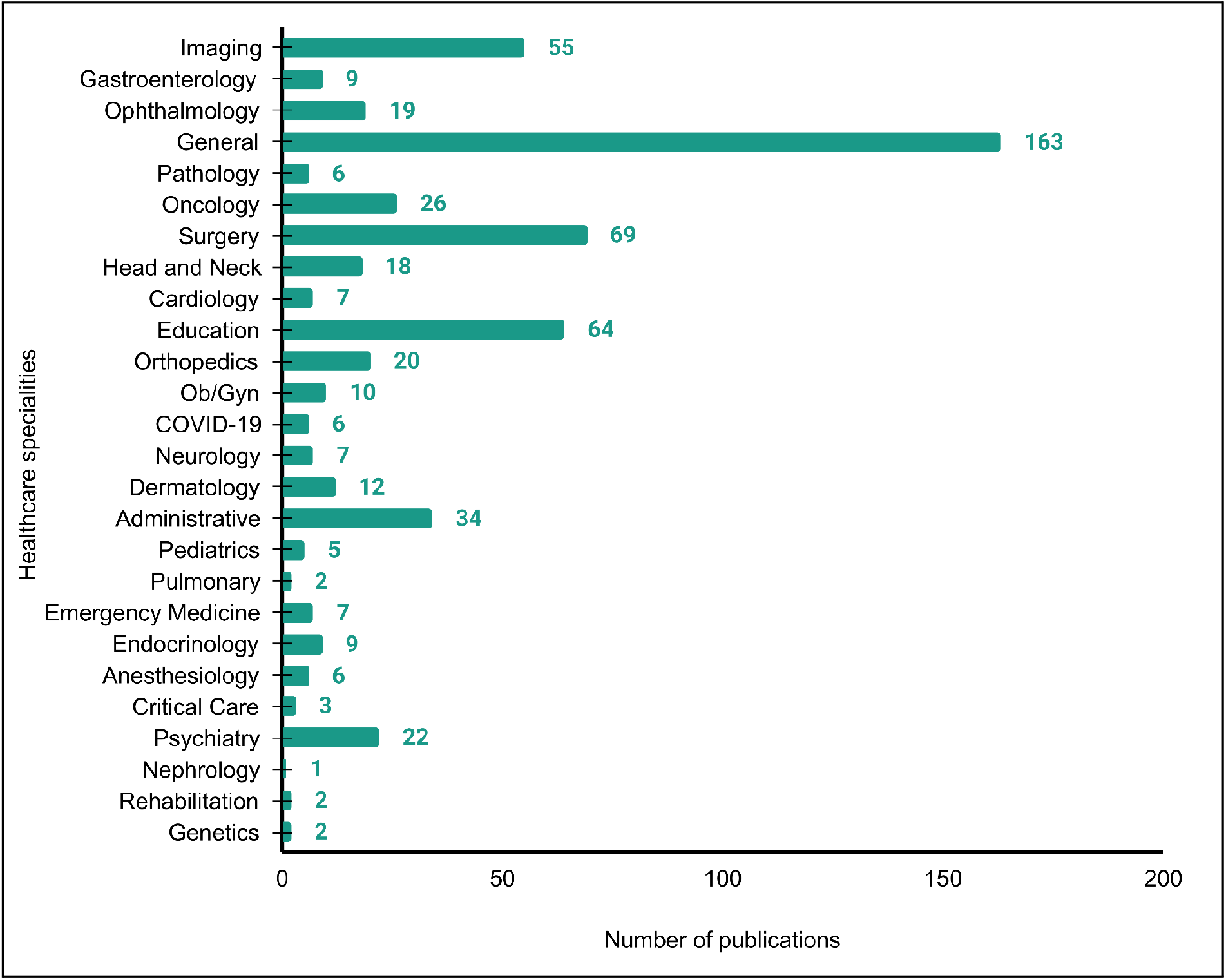
Bar plot of the number of publications based on Large Language Models across healthcare specialties.

## DISCUSSION

The doubling year over year growth rate of AI in healthcare publications continues, with total publication numbers exceeding 23,000 based on our search methodology (**Figure 1**). Similarly, the growth rate of mature publications based on our methodology has also been sustained with mature publications more than doubling in number compared to the year prior (**Figure 1**). With the introduction of ChatGPT and other LLMs since 2022, for the first time significant rise in use of text data(11.6%) and large language models(8.2%) was observed this year. These have historically ranged between 0% to 8.5%[7].

Our methodology was modified from the years prior as it was no longer possible to manually curate such a large number of publications for exclusions, speciality classification or provide subject matter expert viewpoints[7]. Publication in some of the key specialities such as Radiology could possibly have exceeded 10,000. We also believe the trends are reflected in mature publications for each speciality and are adequate to provide substantial information for the purpose of this analysis. We did try various model based classification methods including BERT classifier, LLMs, including prompt engineering LLMs and fine tuning them on our prior years data for the purpose of speciality classification but their results remained suboptimal with classification accuracy parameters ranging from 30-68%.

In 2023, Imaging, GI, Ophthalmology, Oncology, Surgery, Head & Neck, General categories continue to lead the healthcare specialties in the number of mature publications. These specialities represent the healthcare specialities which use images as data type and deep learning models,mostly convolutional neural networks. These healthcare specialties also have a significant number of open source datasets, datathon like competitions and support from their respective societies towards provision of resources to foster AI research[10]. Surgical specialties relative to other medical healthcare specialties tend to outperform in research and publication trends, again primarily related to image based data and application models. COVID-19 publications,as expected, continue to decrease as the pandemic is resolved. Genetics related mature publications also decreased, but the reasons are less clear. Anesthesiology, Critical Care, Nephrology, Rehabilitation continue to have low volumes of publications as they lack image based data for research. Also important to note that in 2021, we observed a drop in publications, which was hypothesized to be due to COVID-19, as in prior publications[7].

Imaging publications continue to focus on various imaging modalities including interpretation of X-rays, CT scans, MRIs and ultrasounds[10–12]. Majority of this research work utilizes deep learning, more specifically and not unexpectedly convolutional neural networks[13,14]. There were a few publications which focus on optimizing image processing and clinician workflows[15]. Most of the Gastroenterology publications continue to focus on trials related to application of deep learning models for endoscopy for diagnosis or to measure treatment response[16,17]. Surgical publications including those for Head and Neck, Orthopedics are focused on image analysis using deep learning to guide surgical diagnosis or decision making.Its interesting to observe a significant number of dentistry research work related to use of image interpretation of dental X-rays or CT scans[18,19]. Oncology publications use both ML and DL to guide diagnosis and treatment decision making. For oncologic diagnosis,use of AI in various cancer screenings such as for breast,cervical and prostate cancer[20]. For management many of the oncology publications focused on radiotherapy planning and predicting or measuring treatment effects for various cancers[21]. Pathology publications continue to focus on use of AI on whole slide images or cytopathology for diagnosis using deep learning[22,23]. Cardiology publications use multimodal data including EKG, CT angiograms amongst others for diagnosis and prediction of arrhythmias or ischemic heart disease,respectively[24–26].

Similar to the prior years, imaging data emerged as the top data type used this year as well, and deep learning models remained the most utilized model type. A significant increase in the utilization of text data was also observed, which may be attributed to the rapid advancement of large language models (LLMs). This advancement was evident in the distribution of model types across all papers, with LLMs being utilized in 8.2% of cases. Additionally, we have witnessed the development of Large Vision Models (LVMs), indicating progress towards more robust multimodal model development, which is essential for future AI healthcare advancements. Among all mature articles, two articles from the LVM category stood out: one in the medical specialty of Pathology, where the author built a Hybrid Vision Transformer for learning gastric histology[8,27], and the other in the Emergency Medicine, where authors developed multimodal models to facilitate the interpretation of chest radiographs in the Emergency Department[28]. Since the distribution of utilized data types and model types is a primary and key indicator of understanding the diversity in overall AI research development, we hope to see more diverse datasets and models utilized to enhance overall AI in healthcare development

Despite large language models being present prior to November 2022 (**Figure 6**), their use in research related to healthcare was insignificant. Since the release of ChatGPT, there has been a tremendous interest in the use of GPT and large language models in healthcare. Initial publications are related to trials of mostly GPT[29]. These trials have focused primarily in a few defined areas: a.) comparison against clinician performance at various tests, b.) summarization tasks, c.) question and answering tasks for patient or clinician questions or recommendations[30–34]. Hence, publications related to comparison between clinician experts’ performance on proficiency tests such as board examinations were significant[35,36].Almost all these publications though used GPT for comparison and without its modification,i.e.,without further significant prompt engineering or fine tuning.

**Figure 6:**
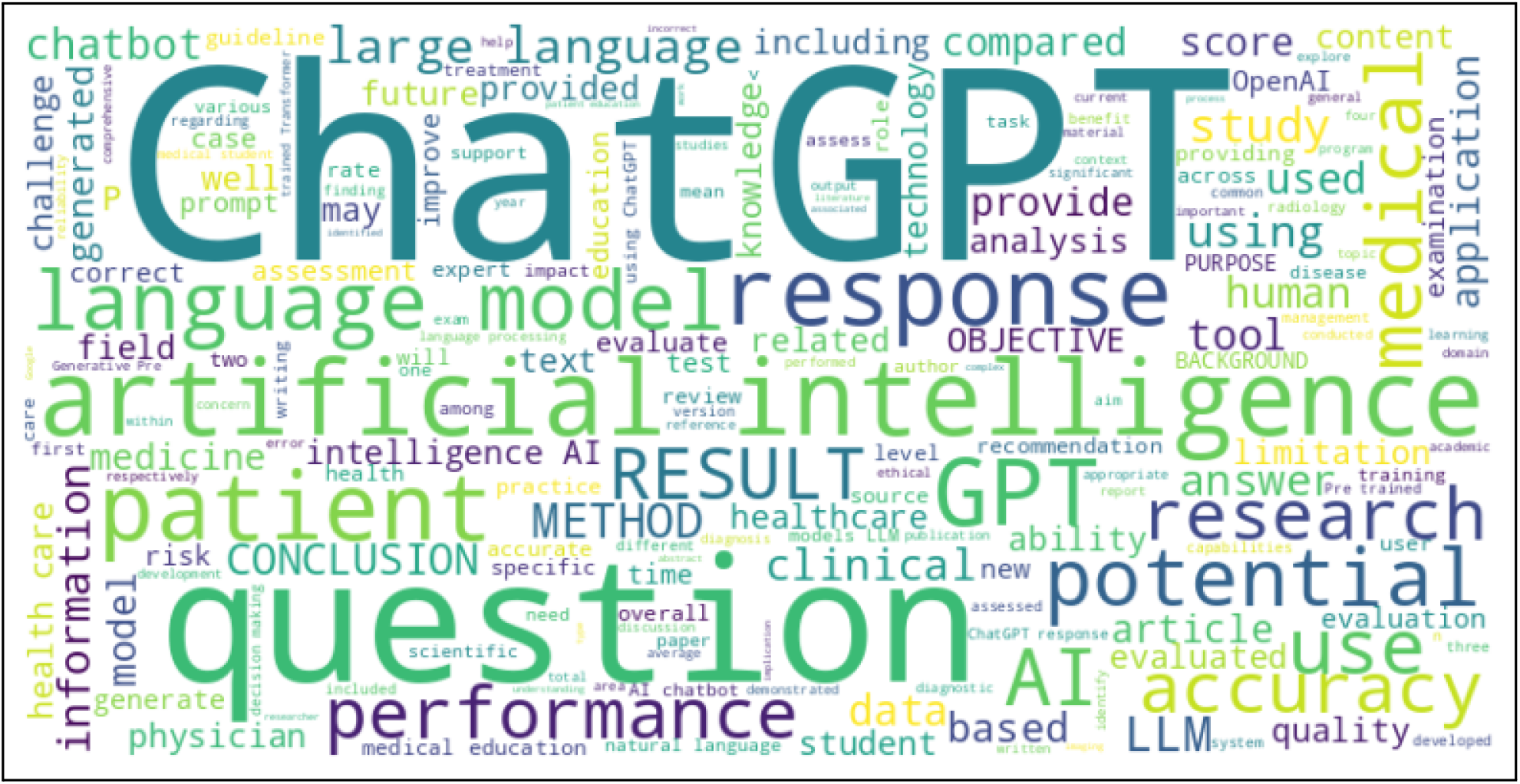
Word cloud of publications on Large Language Models created using abstracts and titles of the publications.

We acknowledge certain limitations of our analysis based on our search or analysis methodology. Search was limited to PubMed and with the restrictions mentioned in the methodology section. It is possible that some of the significant publications might have been missed due to our methodology.In prior years, we have manually excluded many publications from the PubMed search and classified publications into various select healthcare specialities.This year due to the volume of publications(23,306), it was not possible to perform any manual evaluation of all the publications. We did trial various methods including BERT and GPT for automated classification including fine tuning on prior data but the results were not satisfactory. Analysis of mature publications provides a more focused review and a consistent methodology also allows us to provide year over year comparative evaluations[7].

Since, our maturity model is primarily based on the BERT architecture[37], which has limitations such as a lack of understanding of out-of-vocabulary words. BERT’s vocabulary is limited to the tokens it was trained on, so it may struggle with out-of-vocabulary words or rare terms. Therefore, our maturity classification is sensitive if abstracts of publications have used terms that are new for the model. Additionally, the BERT model has a limit in maximum token size of 512, so if the combined length of the title and abstract exceeds 512 tokens, the model may not be able to understand the entire context[37].

We have also evaluated alternate approaches to learn maturity, specifically focusing on methodologies derived from scholarly literature and journal metrics, such as no. of citations, journal impact factors, and journal H-index etc. However, it is important to note that using these metrics to evaluate maturity can be misleading when applied to the analysis of articles from the most recent year due to the unpredictable dynamic trends of these metrics[38–40]. The current BERT-based approach, despite its limitations, remains content-aware, reproducible even with changing factors such as article citation count and journal impact factor, and fine-tunable with new data.

LLM search methodology is limited to search using “LLM” and “GPT” only. This was our preferred method as our trials with other words and sequences yielded a high number of false positive searches.

## CONCLUSION

Exponential growth in research and publications continues in all aspects of AI application in healthcare. Generative AI related publications make a landmark appearance in 2023, bolstering the use of text based data. These publication trends also provide us with early insights into the type of AI and fields of healthcare where AI applications are likely to be implemented or sustained in the future.

## Data Availability

All data produced in the present study are available upon reasonable request to the authors

## DATA AVAILABILITY

Data is available upon request via email to the corresponding author or by contacting us through our website, BrainXAI Research (https://www.brainxai.com/research). Additional related publications and datasets related to AI in Healthcare can be accessed from BrainX Community website(https://www.brainxai.org).

